# Non-falciparum malaria infections are as prevalent as *P. falciparum* among Tanzanian schoolchildren

**DOI:** 10.1101/2022.06.07.22275625

**Authors:** Rachel Sendor, Cedar L. Mitchell, Frank Chacky, Ally Mohamed, Lwidiko E. Mhamilawa, Fabrizio Molteni, Ssanyu Nyinondi, Bilali Kabula, Humphrey Mkali, Erik J. Reaves, Naomi Serbantez, Chonge Kitojo, Twilumba Makene, Thwai Kyaw, Meredith Muller, Alexis Mwanza, Erin L. Eckert, Jonathan B. Parr, Jessica T. Lin, Jonathan J. Juliano, Billy Ngasala

## Abstract

Efforts to achieve malaria elimination need to consider both falciparum and non-falciparum infections. The prevalence and geographic distribution of four *Plasmodium* species were determined by real-time PCR using dried blood spots collected during the 2017 School Malaria Parasitological Survey of eight regions of Tanzania. Among 3,456 schoolchildren, 22% had *P. falciparum*, 24% *P. ovale* spp., 4% *P. malariae*, and 0.3% *P. vivax*. Ninety-one percent of *P. ovale* infections had very low parasite densities, based on amplification at later cycle thresholds. Sixty-four percent of *P. ovale* infections were single-species, and 35% of these were detected in low malaria endemicity regions. *P. malariae* infections were predominantly co-infections with *P. falciparum* (73%). *P. vivax* was largely detected in northern and eastern regions. Overall, 43% of children with *P. falciparum* were co-infected with at least one non-falciparum species. A large, previously under-appreciated burden of *P. ovale* spp. infection exists among Tanzanian schoolchildren.

**Article Summary:** A previously unrecognized burden of non-falciparum malaria infections was detected among Tanzanian schoolchildren in a 2017 cross-sectional study, with *P. ovale* spp. prevalence comparable to *P. falciparum*, and low-level prevalences of *P. malariae* and *P. vivax* detected.

## Background

Sub-Saharan Africa harbors 95% of the global malaria burden^1^. While national surveys conducted by ministries of health throughout Africa regularly assess *Plasmodium falciparum* burden^2^, little is known about the prevalence and geographic distribution of non-falciparum malaria species, including *Plasmodium malariae, Plasmodium vivax*, and *Plasmodium ovale* spp.^3–8^. Although the clinical burden of non-falciparum malaria in sub-Saharan Africa is dwarfed by *P. falciparum*^9^, these species can still elicit symptomatic disease. *P. malariae* has been associated with an increased risk of anemia^10^, as well as clinical complications such as chronic nephrotic syndrome^11,12^. *P. vivax* causes severe anemia, pregnancy-related complications, and mortality following recurrent infections, although infections in sub-Saharan Africa are infrequent^13–15^. Clinical consequences of *P. ovale* spp. infection are least well-described, though well-described in travelers, and have been associated with severe infection in case reports^16^.

Recent findings suggest declining *P. falciparum* prevalence in East Africa may be associated with increases in non-falciparum infections^17–20^. However, comprehensive surveys of non-falciparum malaria in sub-Saharan Africa are infrequent because detection of these species remains challenging^11,17^. Existing field diagnostic methods such as microscopy and pan-*Plasmodium* spp. lactate dehydrogenase (LDH) or histidine-rich protein 2 (HRP2)-based rapid diagnostic tests (RDTs) lack sensitivity to detect non-falciparum species^11,17^. Non-falciparum parasite densities are often low, and the majority of those infected may not seek care. Mixed-infections with *P. falciparum* can also complicate detection of concurrent infection by non-falciparum species^3,17^. Molecular detection methods can sensitively detect non-falciparum species, but remain largely confined to research use.

In Tanzania, malaria burden is high, with the country accounting for 4.1% of malaria deaths globally in 2020^1^. While approximately 93% of the mainland Tanzanian population remains at risk of infection, transmission throughout the country is highly heterogeneous^21^. Malaria transmission patterns are largely driven by geographic features of the country: across the arid highlands and in urban centers, malaria transmission is low, unstable, and seasonal; in southern, northern, and northwestern areas, transmission is moderate and varies seasonally; and along the coastal, lake, and southern regions, malaria transmission is high and perennial^21,22^.

Decades of concentrated malaria control interventions have helped lower the national prevalence from 18% in 2008 to 7% in 2017^23^. The vast majority of malaria cases reported in Tanzania are attributed to *P. falciparum*^9,21^, although recent studies have also identified evidence of *P. malariae, P. vivax*, and *P. ovale* transmission^4,18,24,25^. Given widespread use of *P. falciparum-*specific HRP2-based RDTs for malaria diagnosis, the propensity for missed detection or misclassification of non-falciparum species in Tanzania is high, and large-scale, geographically representative studies to assess spatial distributions of these species are lacking.

This study deployed molecular methods in the context of a national survey to comprehensively characterize non-falciparum malaria epidemiology among schoolchildren in Tanzania. We demonstrate a high prevalence of *P. ovale* spp. infection, on par with *P. falciparum* prevalence. These infections vary geographically and do not always correlate with *P. falciparum* prevalence in a given region, with many *P. ovale* spp. infections detected in regions considered at low epidemiological risk for malaria. *P. malariae* was also broadly detected, whereas *P. vivax* was detected in limited geographical regions.

## Methods

### Study Design

The 2017 School Malaria Parasitological Survey (SMPS) was a cross-sectional study conducted in 2017 among children aged 5–16 years enrolled in public primary schools in mainland Tanzania. Methods for site selection and survey design mirrored the 2015 SMPS and have been previously summarized in detail^22^. Regions included in the study were selected through a multi-stage sampling scheme to maintain geographic representation and reflect the heterogeneity of malaria transmission across Tanzania^22,26^. The number of schools randomly selected per region was proportional to each region’s respective population^22,26^. Within each school, an average of 100 students were randomly selected for screening, and consented students were interviewed for demographic and clinical characteristics, had a malaria RDT performed, and provided a dried blood spot (DBS) sample^22,26^. Surveys largely coincided with each region’s rainy season. Among students who provided a DBS, a stratified random sample was selected to construct a sub-population for non-falciparum malaria testing, with students sampled in equal proportion to the regional proportion of students within the broader survey population to maintain representativeness.

Malaria detection was conducted using CareStart™ Malaria Pf/PAN (HRP2/pLDH) Ag Combo RDTs (Product code: RMRM 02571; Access Bio, Inc., Somerset, NJ, USA) detecting *P. falciparum*-specific HRP2 and pan-pLDH antigens; RDTs were considered positive if either band was positive. Schools and councils were grouped into epidemiological strata of malaria transmission risk based on *P. falciparum* prevalences in children estimated from the 2014-15 Tanzania SMPS^22,26^. *P. falciparum* prevalence was considered “very low” if <5%, “low” 5 to <10%, “moderate” 10 to <50%, and “high” ≥50%^22,26^. Dried blood spots (DBS) were collected on Whatman filter paper (Whatman plc, Cytiva, Buckinghamshire, UK) and shipped to the University of North Carolina, Chapel Hill for molecular testing.

### Molecular Detection

DNA was extracted from three 6mm DBS punches using Chelex methods^27^, and samples underwent real-time PCR targeting the 18S ribosomal subunit for malaria testing, according to previously published protocols^28^ (**Supplementary Table 1**). Assays for each *Plasmodium* species (*falciparum, ovale* spp., *malariae*, and *vivax*) were run independently with appropriate controls. Positive controls for *P. falciparum* detection were made using whole blood and 3D7 cultured parasites (MRA-102; BEI Resources) to create mocked DBS, with DNA extracted using Chelex^27^ and serially diluted. Positive controls for non-falciparum species were based on standard dilutions of plasmid DNA from MR4 (MRA-180, MRA-179, MRA-178; BEI Resources), with semi-quantitative parasitemia estimated using six 18S rRNA gene copies per parasite^28^. *P. malariae* and *P. falciparum* assays were run for 40 cycles. *P. ovale* spp. and *P. vivax* assays were run for 45 cycles to enable detection of very-low density infections^28^. This approach was previously validated using 390 negative controls (comprising water [n=22] and human DNA [n=368]) and >170 positive controls of decreasing parasite densities per non-falciparum species; no false-positives were detected^28^. Assay specificity was also assessed previously by testing against other *Plasmodium* species^29^. In addition, the UNC laboratory participates in the World Health Organization malaria molecular quality assurance scheme, identifying and determining species of *Plasmodium* parasites in blinded samples every six months, with consistent high marks for assay performance across species. In the present study, among 20 negative controls per each species-specific assay, no false positive amplification was detected (**Supplementary Table 2**). A subset of positive *P. ovale* samples underwent further real-time PCR testing to distinguish *wallikeri* and *curtisi* species^30,31^. To evaluate potential bias due to differences in PCR cycles between species, a sensitivity analysis was conducted in a random sample of 750 students stratified by malaria transmission intensity, detecting *P. falciparum* and *P. malariae* infections by semi-quantitative, real-time PCR of the 18S RNA gene performed to 45 cycles.

### Analysis

Malaria species-specific prevalences were calculated overall, and by single vs. mixed-species infections. Prevalences were not adjusted with sampling weights as samples for the non-falciparum analysis population were selected randomly and in equal proportion to the broader survey sample.

Descriptive statistics were computed for student-level characteristics by *Plasmodium* species. Differences between *P. falciparum* and non-falciparum species mono-infections were tested using Pearson’s chi-square analysis and Kruskal-Wallis rank sum testing in the presence of non-normality; Fisher’s exact test was applied where cell counts were <5. Similar analyses were performed comparing PCR–positive vs. PCR–negative students by species. Missing data were excluded from statistical analysis.

### Spatial Mapping

Regional variation in the prevalence of each species was assessed through geospatial mapping of infections by council and region. Number of infections and number of students tested were aggregated by council, and prevalence estimates by council were estimated and mapped.

Scaled prevalence estimates were calculated by taking the proportion of each council’s prevalence out of the highest council prevalence for each species, as follows:

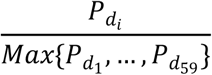

where *P* is the prevalence for a given council, *d*_*i*_.

Differences between scaled non-falciparum prevalence and scaled *P. falciparum* prevalence for each council were calculated and mapped to determine differential scaled prevalences. This method allowed comparison of prevalence estimates between each non-falciparum species with *P. falciparum*, while accounting for differences in the absolute burden of each species.

Analyses and mapping were performed using R (version 4.0.2), including *eulerr*^32^ and *sf* (version 0.9-7)^33^ packages. Shapefiles were sourced from GADM.org and elevation measurements were collected from the Shuttle Radar Topography Mission. Informed consent was obtained from students and their legal guardians prior to survey or blood sample collection and ethical clearance was obtained from the Tanzania National Institute for Medical Research. Analysis of de-identified samples was approved by the Institutional Review Board of the University of North Carolina, Chapel Hill (IRB #19-1495).

## Results

### Study Population

A total of 3,456 students from 180 schools across eight geographically-representative regions were selected for non-falciparum testing from among 17,131 students in the SMPS who had a DBS sample available. No significant differences were detected between the sub-population for non-falciparum testing and the SMPS DBS population (**Supplementary Table 3**). Median (IQR) student age in the non-falciparum analysis population was 11 (9–13) years, with an approximately equal distribution of males and females (51% vs. 49%). Malaria dual-antigen RDTs were positive in 20% of students. The majority of students attended schools in regions classified as ‘high’ (51%) or ‘moderate’ (13%) malaria transmission risk (**Table 1**).

**Table 1.**
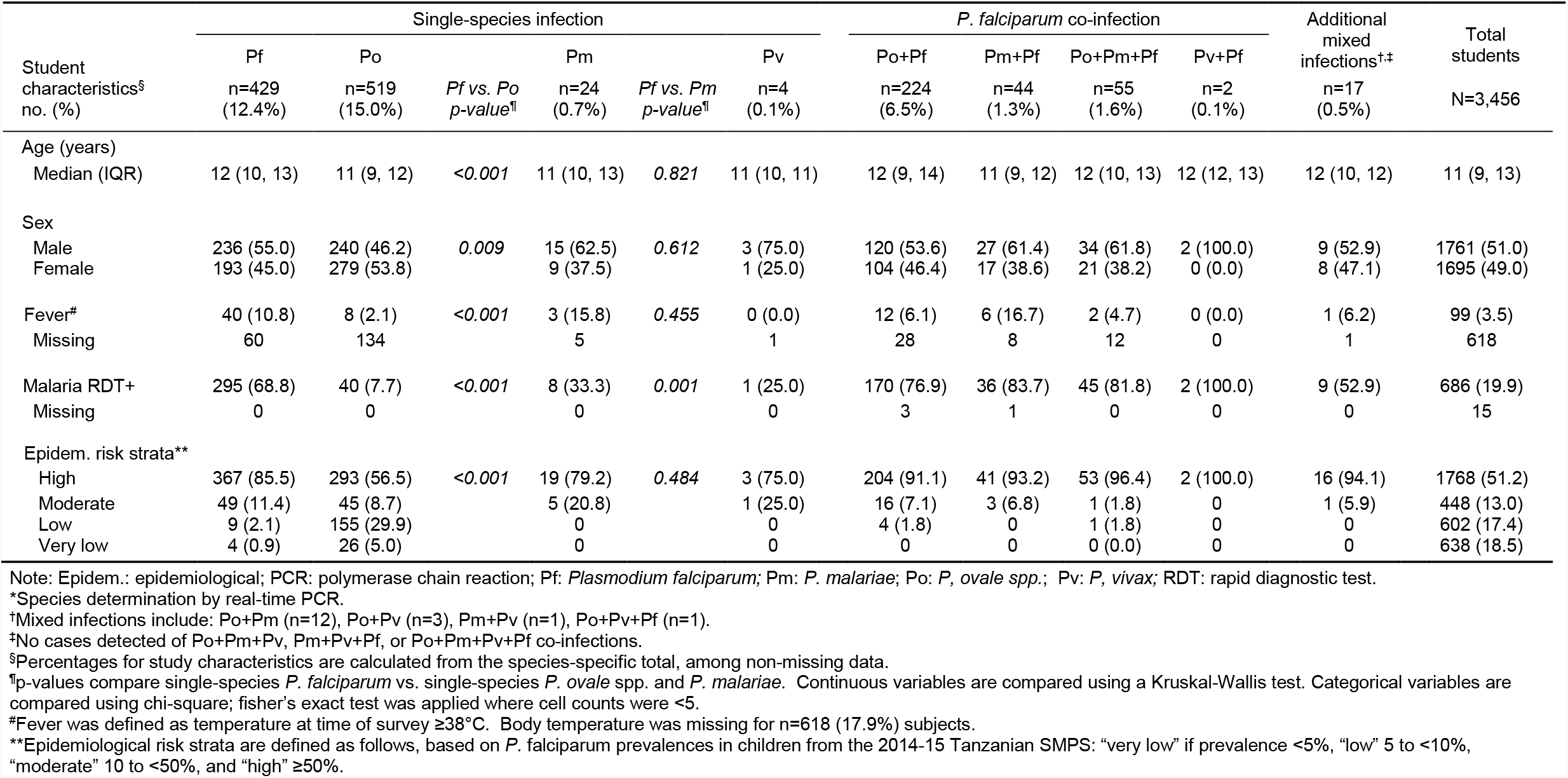
Student infection characteristics by malaria species*

### Species Prevalence by PCR

*P. falciparum* infection was identified in 22% (n=755; 95% CI: 21%–23%) of students, *P. ovale* spp. in 24% (n=814; 95% CI: 22%–25%), *P. malariae* in 4% (n=136; 95% CI: 3%–5%), and *P. vivax* in 0.3% (n=11; 95% CI: 0.2%–0.6%), including single and mixed-species infections (**Supplemental Table 4**).

The majority of *P. ovale* spp. infections identified were single-species (64%, n=519), with 28% (n=224) mixed with *P. falciparum* only. Conversely, most *P. malariae* infections were co-infected with both *P. ovale* spp. and *P. falciparum* (40%, n=55), or with *P. falciparum* only (32%, n=44) [**Figure 1**]. Thirty-six percent (n=4) of *P. vivax* infections were single-species. Forty-three percent (n=326) of *P. falciparum* infections were co-infected with at least one non-falciparum species.

**Figure 1.**
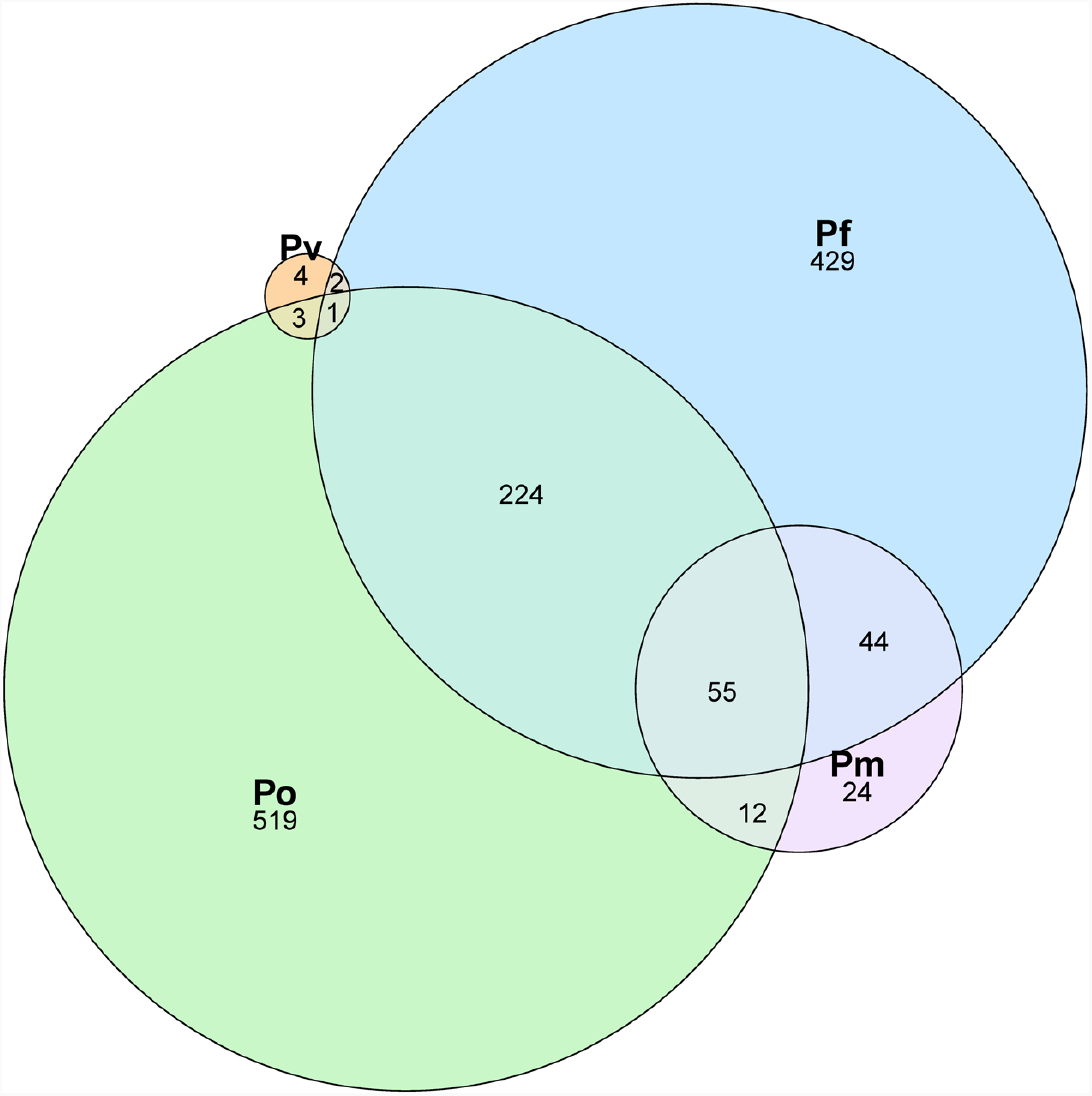
Distribution of malaria infection by species among Tanzanian schoolchildren Note: Pf: *P. falciparum;* Pm: *P. malariae*; Po: *P. ovale* spp.; Pv: *P. vivax*. N=1 *Pv + Pm* co-infection is not depicted. Prevalence estimates by species are: *P. falciparum*: 21.8% (n=755; 95% CI: 20.5% – 23.3%); *P. ovale* spp.: 23.6% (n=814; 95% CI: 22.2% – 25.0%); *P. malariae*: 3.9% (n=136; 95% CI: 3.3% – 4.6%); *P. vivax*: 0.3% (n=11; 95% CI: 0.2% – 0.6%).

To evaluate the impact of different PCR cycles between assays, a sensitivity analysis testing *P. falciparum* and *P. malariae* within 45 PCR cycles was performed. A 25% (95% CI: 21%–29%) prevalence of *P. falciparum* and 3% (95% CI: 2%–5%) prevalence of *P. malariae* was observed, weighted to the distribution of students in the total non-falciparum population by transmission intensity strata. Within this subset, 0.5% (n=4) of *P. falciparum* and 0.3% (n=2) of *P. malariae* infections were detected at Cts between 40 to 45 (**Supplemental Table 5**). Thus, >99% of *P. falciparum* and *P. malariae* infections were detected by the primary 40-cycle assays utilized in the study.

Differences in subject-level characteristics between species are depicted in **Table 1** (and **Supplemental Table 6**). Compared to *P. falciparum, P. ovale* spp. mono-infections were detected more frequently in slightly younger (median 11 vs. 12 years; p<0.001) and female (54% vs. 45%; p=0.009) students. In terms of sensitivity by RDT compared to PCR, 8% (n=40) of *P. ovale* spp. mono-infections were RDT-positive for any band, whereas 33% (n=8) of *P. malariae* and 69% (n=295) of *P. falciparum* mono-infections were RDT-positive. Non-falciparum-*P. falciparum* co-infections were positive by RDT in 78% (n=253/325) of infections detected by PCR.

Though only 3% (n=13) of *P. falciparum* mono-infections and no *P. malariae* or *P. vivax* mono-infections were detected in low-risk strata, 35% (n=181) of *P. ovale* spp. mono-infections occurred in regions considered to have ‘low’ or ‘very low’ epidemiological risk for malaria. ‘High” epidemiologic risk strata harbored the majority of mono-infections across all four malaria species, as well as mixed-infections with *P. falciparum*.

### Parasite Density

Malaria parasitemia estimated by semi-quantitative PCR was low across non-falciparum species (**Figure 2**). The median (IQR; min–max) parasite densities for *P. ovale* spp. and *P. malariae* were comparable (1.8 [0.3 – 6.2; 0.02 – 42,149]) *ovale* parasites (p)/μL and 2.9 [0.7 – 13.7; 0.1 – 304] *malariae* p/μL, respectively). *P. vivax* parasite densities were an order of magnitude lower, around 0.1 (0.1 – 0.2; 0.02 – 2.0) *P. vivax* p/μL. Whereas a small proportion of *P. malariae* infections had a parasite density >50 p/μL (12%, n=16), this was rarely observed for *P. ovale* spp. (1%, n=11), and never the case for *P. vivax*, where the highest parasite density detected was 2.0 p/μL. Median (IQR; min-max) *P. falciparum* parasite density was also low at 13.1 (2.6 – 55.9; 0.1 – 8,248) p/μL, although 27% (n=203) of *P. falciparum* cases had a parasite density >50 p/μL, and 3% (n=24) >500 p/μL.

**Figure 2.**
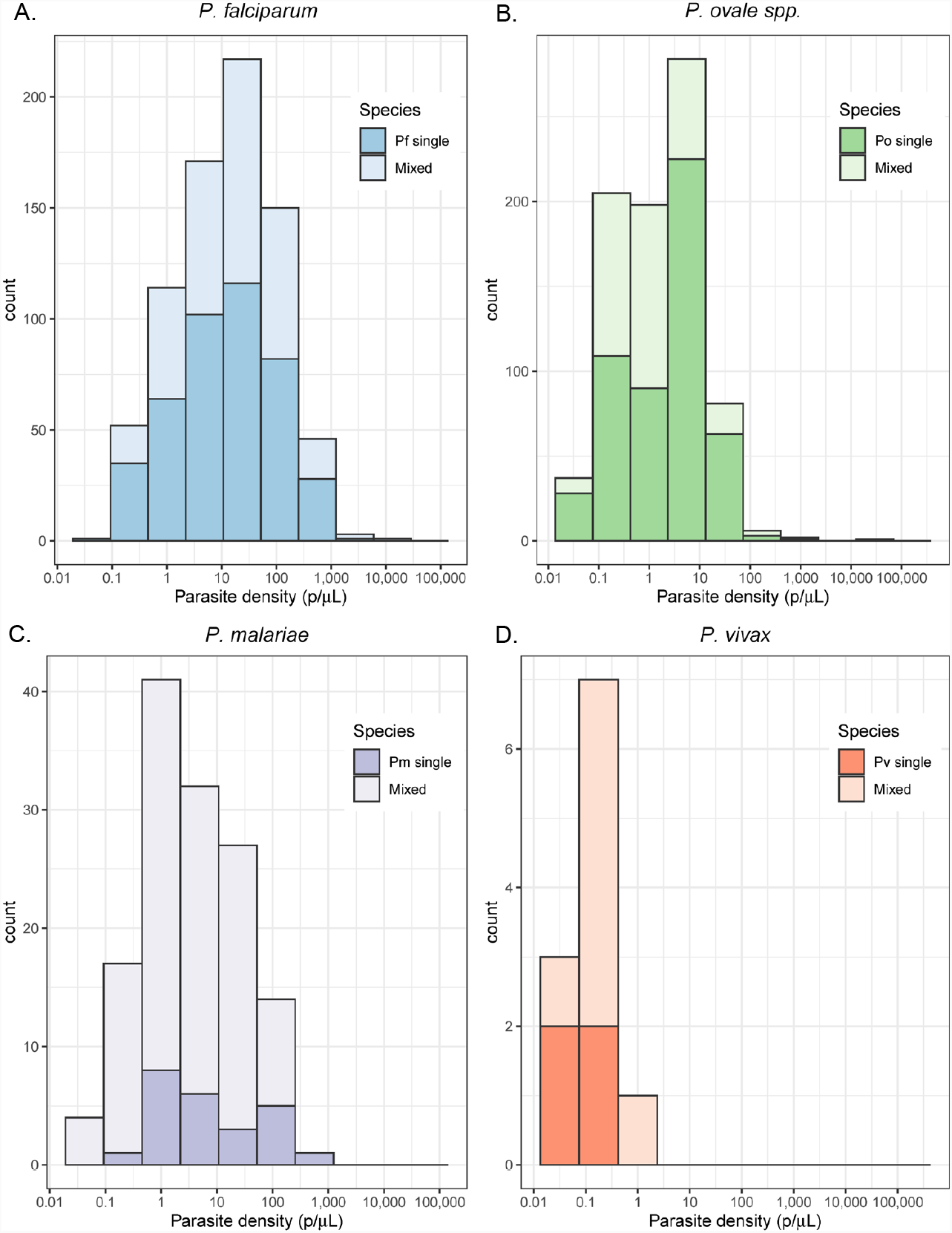
Estimated parasite density (p/μL) distributions by malaria species Note: Parasite densities displayed on log scale. Mixed infections include *P. falciparum* and non-falciparum co-infections. *P. ovale* and *P. vivax* parasite densities were detected by including isolates amplifying at later PCR cycle thresholds. **A)** *P. falciparum* median (IQR) parasite density: (single-species [n=429]: 11.4 [2.5-54.7] p/μL; mixed [n=326]: 16.5 [3.5-56.9] p/μL; p=0.117). **B)** *P. ovale* spp. median (IQR) parasite density: (single-species [n=519]: 3.4 [0.3-7.5] p/μL; mixed [n=295]: 0.8 [0.3-2.8] p/μL; p<0.001). **C)** *P. malariae* median (IQR) parasite density: (single-species [n=24]: 4.0 [0.9-40.9] p/μL; mixed [n=112]: 2.8 [0.6-13.5] p/μL; p=0.169). **D)** *P. vivax* median (IQR) parasite density: (single-species [n=4]: 0.1 [0.1-0.2] p/μL; mixed [n=7] 0.2 [0.1-0.2] p/μL; p=0.571).

Median (IQR) parasite density among *P. ovale* spp. mixed-infections was lower than among *P. ovale* spp. mono-infections (0.8 [0.3-2.8] *ovale* p/μL vs. 3.4 [0.3-7.5] p/μL; p<0.001), whereas densities were similar between single and mixed-species infections among all other malaria species (**Figure 2**).

### Plasmodium ovale Species Determination

Among 814 samples positive for *P. ovale* spp. infection, 60 (7%) of the highest parasite density samples were selected for real-time PCR detection of *P. ovale wallikeri* and *P. ovale curtisi* species. Species determination was successful in 35% (n=21) of samples, with *P. ovale curtisi* detected in 17 samples and *P. ovale wallikeri* in 9. Five students were positive for both *P. ovale curtisi* and *P. ovale wallikeri* infection. Further characterization by demographic and clinical characteristics was not performed due to limited sample size.

### Geographic Distribution

*P. ovale* spp. were detected across all eight regions sampled in the study, indicating widespread distribution across Tanzania (**Table 2, Figure 3**). Prevalence of *P. ovale* spp. was highest within the northern Kagera region (34%, n=273) and the central region of Tabora (17%, n=139). *P. ovale curtisi* infections were detected in six of eight study regions (all but Arusha and Rukwa), and *P. ovale wallikeri* was detected in five (Kagera, Mara, Tabora, Tanga, and Iringa). High prevalence of *P. malariae* was also detected in Kagera (29%, n=39), similar to *P. ovale* spp., and in the southernmost Mtwara region (28%, n=38). *P. vivax* infections were predominantly distributed along the northern-western border of Tanzania in Kagera (55%, n=6), with select, isolated cases of single and mixed-species *P. vivax* infections also detected in southern and eastern regions. The Arusha region in the northeast and Iringa in the southwest did not have any cases of *P. malaria*e or *P. vivax* infections, and had the lowest frequencies of *P. ovale* spp. (3%, n=23; 4%, n=30) and *P. falciparum* infections (0.4%, n=3; 0.1%, n=1).

**Table 2.** Student infections by *Plasmodium* species and school characteristics across Tanzania

| School characteristics<sup>†</sup><br>no. (%) | *Plasmodium* infections*<br>(single and mixed species) |  |  |  | Total students<br>N=3,456 |
| --- | --- | --- | --- | --- | --- |
|  | Pf<br>n=755 | Po<br>n=814 | Pm<br>n=136 | Pv<br>n=11 |  |
| Elevation (m) |  |  |  |  |  |
| Median (IQR) | 1182<br>(506, 1370) | 1225<br>(1124, 1427) | 1167<br>(320, 1370) | 1333<br>(1100, 1398) | 1230<br>(1058, 1467) |
| Min, Max | 54, 1901 | 47, 2167 | 54, 1677 | 184, 1467 | 34, 2167 |
| <1500 | 707 (26.5) | 693 (26.0) | 129 (4.8) | 11 (0.4) | 2667 (100) |
| ≥1500 | 48 (6.1) | 121 (15.3) | 7 (0.9) | 0 | 789 (100) |
| Region<sup>‡</sup> |  |  |  |  |  |
| Arusha | 3 (0.5) | 23 (4.2) | 0 | 0 | 552 (100) |
| Iringa | 1 (0.3) | 30 (9.4) | 0 | 0 | 320 (100) |
| Kagera | 196 (31.7) | 273 (44.1) | 39 (6.3) | 6 (1.0) | 619 (100) |
| Mara | 157 (34.7) | 102 (22.6) | 19 (4.2) | 2 (0.4) | 452 (100) |
| Mtwara | 146 (47.6) | 62 (20.2) | 38 (12.4) | 1 (0.3) | 307 (100) |
| Rukwa | 49 (16.3) | 118 (39.2) | 6 (2.0) | 0 | 301 (100) |
| Tabora | 122 (29.5) | 139 (33.7) | 25 (6.1) | 0 | 413 (100) |
| Tanga | 81 (16.5) | 67 (13.6) | 9 (1.8) | 2 (0.4) | 492 (100) |

Note: Pf: *Plasmodium falciparum*; Pm: *P. malariae*; Po: *P. ovale* spp.; Pv: *P. vivax*.
\*Infection totals represent student-level infection prevalences. Total infections are presented, encompassing single and mixed-species infections.
<sup>†</sup>Row percentages are calculated; percentages are not mutually exclusive given co-infections are included in species-specific columns.
<sup>‡</sup>Regions correspond to mapped regions in Figure 3.

**Figure 3.**
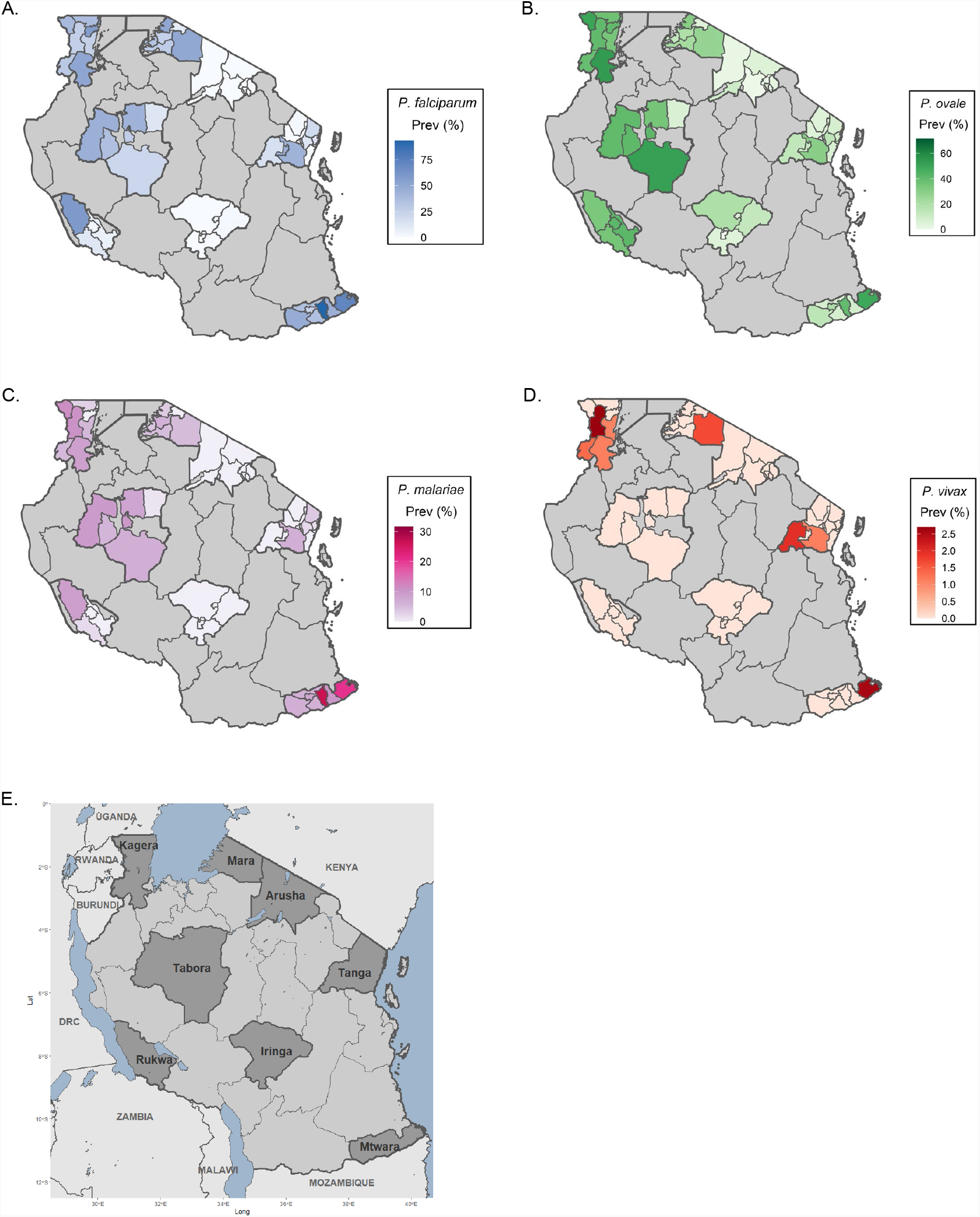
School council-level malaria prevalence distributions by species.

Malaria infections across all species were predominantly detected at elevations <1500 meters (m), including 85% (n=693) of *P. ovale* spp., 94% (n=707) of *P. falciparum*, 95% (n=129) of *P. malariae*, and 100% (n=11) of *P. vivax* infections (**Table 2**). In comparison, 77% (n=2667) of students enrolled in the study were derived from schools at elevations <1500m.

Among those enrolled at higher elevations, *P. ovale* spp. infections were detected most frequently at 15% (n=121), compared to 6% (n=48) of students with *P. falciparum*, 1% (n=7) *P. malariae*, and 0 *P. vivax*.

Comparing scaled prevalence estimates for non-falciparum species with *P. falciparum*, we identified areas where prevalences were higher than expected for *P. ovale* spp. and *P. malariae*, based on *P. falciparum* burden (**Figure 4**); *P. vivax* infections were too infrequent for comparison. In the southern and south-western highlands and north-western lake regions (Iringa, Rukwa, Tabora, and Kagera) scaled *P. ovale* spp. prevalences were higher relative to *P. falciparum*. Scaled prevalence of *P. malariae* was notably higher than *P. falciparum* in the Karagwe council in Kagera and the Mtwara municipal council in Mtwara. In most other areas, scaled prevalence of *P. malariae* was similar or lower to *P. falciparum* prevalence.

**Figure 4.**
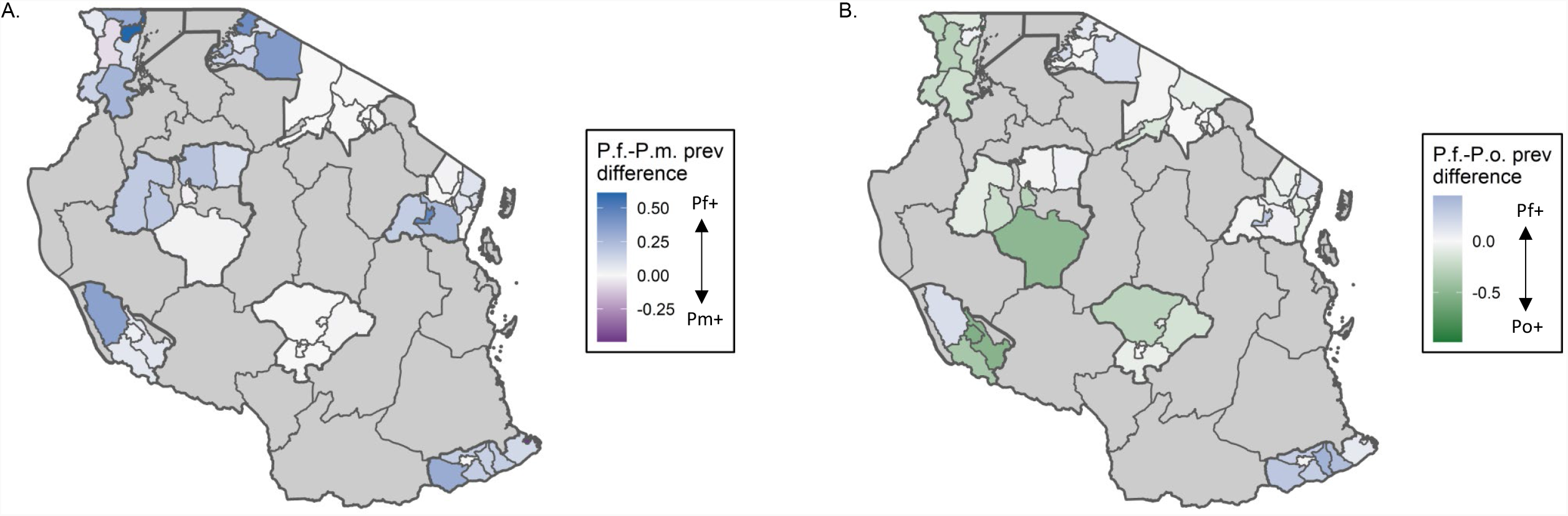
Differential scaled prevalences of *P. malariae* and *P. ovale* spp. compared to *P. falciparum* at the school council-level Note: scaled prevalence of *P*.*f*–*P*.*v*. is not depicted given the low number of *P. vivax* infections biasing the scaled measure. **A)** Blue shading indicates councils where *P. falciparum* scaled prevalence is greater than *P. malariae* scaled prevalence in that council. Purple indicates regions where *P. malariae* scaled prevalence is greater. **B)** Blue shading indicates councils where *P. falciparum* scaled prevalence is greater than *P. ovale* spp. scaled prevalence in that council. Green indicates regions where *P. ovale* spp. scaled prevalence is greater.

## Discussion

This study represents the largest nationally representative molecular survey of non-falciparum malaria epidemiology across Tanzania to our knowledge. Real-time PCR was employed to estimate non-falciparum infection prevalences within school-aged children in eight regions of the country selected to maintain geographic diversity and heterogeneity of malaria transmission risk. One-quarter of schoolchildren (24%) harbored *P. ovale* spp. parasites, comparable to the detected *P. falciparum* prevalence (22%) in the population, with more than half (64%) constituting single-species infections. *P. malariae* was observed in 4% of students, with the majority co-infected with other malaria species. *P. vivax* infections were rare but present at a 0.3% prevalence.

The high prevalence of *P. ovale* spp. may be attributed to several factors. First, we extended the cycling time of our *P. ovale* PCR assay to 45 cycles to enable detection of low-density infections, which comprised 91% of all *P. ovale* spp. infections identified. There is precedent for this approach^17,25,31^, as low density parasitemia is characteristic of *P. ovale* spp. infections, making them challenging to detect. The use of 40-cycle *P. ovale* PCR assays yielded a lower 0.8% prevalence estimate in our prior work in the Democratic Republic of Congo (DRC)^5^. The prevalence of *P. ovale* spp. infections positive at <40 cycles in the present study was 9% (n=75), confirming the majority of infections occurred at very-low parasite densities. Second, many large-scale molecular surveys of non-falciparum malaria have focused on adults or all-age cohorts, whereas school-aged children are increasingly recognized as the main contributors to asymptomatic and infectious malaria reservoirs^34–36^. Finally, the high prevalence of *P. ovale* spp. in our study may reflect increasing *P. ovale* spp. transmission in the face of malaria control efforts targeting *P. falciparum*. Increasing or persistent transmission of *P. ovale* and *P. malariae* amid *P. falciparum* decline has been observed in molecular surveys from Tanzania and nearby Kenya and Uganda, including in symptomatic cases^17,18,24,37^. It’s unclear whether this is due to hypnozoite-induced relapses from *P. ovale* spp. infections not treated by artemisinin-based combination therapies (ACTs), day-biting or outdoor vectors that evade bed nets, or other factors.

In contrast to findings from other studies^11,37–40^, *P. ovale* spp. more commonly occurred as single-species infections compared to the other non-falciparum species, though this finding may be due in part to the increased sensitivity of the *P. ovale* PCR assay employed. These single-species infections were rarely detected by RDTs, rendering them more difficult to detect and treat. Single-species *P. ovale* infections were also largely the only species identified within regions categorized as low risk for malaria transmission, suggesting an unexpected transmission risk in areas where prevention measures may be less frequently used and *P. falciparum* risk is not a large concern. Our scaled differential prevalence map similarly highlights several councils where *P. ovale* spp. and *P. malariae* prevalence are proportionately higher than would be expected based on *P. falciparum* burden. Taken together, these characteristics indicate a hidden burden of *P. ovale* spp. in the country.

Detection of *P. vivax* in this study is notable given the control challenges this species poses. Infections were predominately detected in the northwest/Lake zone regions of Tanzania and in the east, where several studies have also observed low-level *P. vivax* prevalences^4,24,41^. The *P. malariae* prevalence of 4% detected in this study aligns with recent research in the region identifying a low prevalence of infection (2.5% in Malawi, 4.1% in DRC, 3.3% and 5.3% [symptomatic and asymptomatic] in western Kenya)^12,28,40^. Estimated parasite densities were low across all non-falciparum species, as expected. *P. falciparum* parasite densities were relatively low as well (median 13.1 p/μL), likely due to the survey of a predominantly asymptomatic population. Additionally, prevalence mapping confirmed low or non-existent prevalence of non-falciparum malaria within the northern highlands of Arusha and southern highlands/midlands of Iringa.

This study has several limitations. The use of different PCR cycling times by species introduces ascertainment bias. Because *P. malariae* and *P. falciparum* assays were run at 40 rather than 45 cycles, their relative prevalence compared to *P. ovale* spp. may be underestimated. However, a sensitivity analysis to quantify this bias indicated we would expect to detect only an additional 0.5% of *P. falciparum* and 0.3% of *P. malariae* infections if testing to 45 cycles, suggesting minimal underestimation of reported *P. falciparum* and *P. malariae* prevalences, and no meaningful impact on overall conclusions. Weighting sensitivity analysis results to the total study population yielded a *P. falciparum* prevalence of 25% if run to 45 cycles, compared to the observed prevalence of 22%. Additionally, this study did not sample all geographic regions in Tanzania, and findings cannot be extrapolated to other age groups with differing malaria risk profiles. School-based sampling likely underestimates prevalence of infection from symptomatic or severe malaria infections in school-aged children, as children may be absent due to illness. Lastly, the cross-sectional survey design reveals little about clinical implications of prevalent non-falciparum infections, especially given substantial, non-random missingness in fever data, or to what extent these infections represent chronic carriage versus transient parasitemia.

The overall high prevalence and broad geographic distribution of *P. ovale* spp. and to a lesser extent, *P. malariae*, with more focal distribution of *P. vivax*, detected in this study underscore an urgent need to understand clinical burden and transmission patterns of these species to inform malaria control programs in Tanzania. Current Tanzanian treatment protocols do not regularly treat for hypnozoite liver-stage *P. ovale* spp. infection, and relapses are expected after blood-stage clearance by ACTs^42^. With accumulating evidence of a previously unappreciated non-falciparum malaria burden present in sub-Saharan Africa^39^, new detection and treatment strategies may be required for continued progress toward malaria control and elimination.

## Supporting information

STROBE Checklist

## Data Availability

Data are available upon reasonable request to the authors.

## Acknowledgments

We thank the 2017 SMPS study administrators and staff for their tireless work implementing the survey and we are grateful for the participation of all individuals in the study. The following reagents were obtained through BEI Resources, NIAID, NIH: Diagnostic Plasmid Containing the Small Subunit Ribosomal RNA Gene (18S) from *Plasmodium malariae*, MRA-179, *Plasmodium ovale*, MRA-180, and *Plasmodium vivax*, MRA-178, contributed by Peter A. Zimmerman. The following reagent was obtained through BEI Resources, NIAID, NIH: *Plasmodium falciparum*, Strain 3D7, MRA-102, contributed by Daniel J. Carucci.

## Disclaimer

The findings and conclusions in this report are those of the author(s) and do not necessarily represent the official position of the U.S. Centers for Disease Control and Prevention, the President’s Malaria Initiative via the U.S. Agency for International Development, or other employing organizations or sources of funding.

## Data Availability

Data are available upon reasonable request to authors.

