## Supplemental Tables 1-6 for "Non-falciparum malaria infections are as prevalent as *P. falciparum* among Tanzanian schoolchildren"

**SUPPLEMENT**

Supplemental Table 1: PCR assay primers and probes by *Plasmodium* species

| *Plasmodium* species | Factor | Final molarity | Sequence (5' to 3') |
|---|---|---|---|
| *malariae* | Forward Primer | 300 nM | AGTTAAGGGAGTGAAGACGATCAGA |
| | Reverse Primer | 300 nM | CAACCCAAAGACTTTGATTTCTCATAA |
| | Probe | 200 nM | FAM/ATGAGTGTTTCTTTTAGATAGC/MGBNFQ |
| *ovale* spp. | Forward Primer | 400 nM | CCRACTAGGTTTTGGATGAAAVRTTTTT |
| | Reverse Primer | 400 nM | AACCCAAAGACTTTGATTTCTCATAA |
| | Probe | 200 nM | VIC/CRAAAGGAATTYTCTTATT/MGBNFQ |
| *vivax* | Forward Primer | 400 nM | ACGCTTCTAGCTTAATCCACATAACT |
| | Reverse Primer | 400 nM | ATTTACTCAAAGTAACAAGGACTTCCAAGC |
| | Probe | 200 nM | FAM/TTCGTATCG/ZEN/ACTTTGTGCGCATTTTGC/3IABkFQ |
| *falciparum* | Forward Primer | 300 nM | ATTGCTTTTGAGAGGTTTTGTTACTTT |
| | Reverse Primer | 300 nM | GCTGTAGTATTCAAACACAATGAACTCAA |
| | Probe | 200 nM | FAM/CATAACAGACGGGTAGTCAT |
| *ovale curtisi* | Forward Primer | 300 nM | TTTTGAAGAATACATTAGGATACAATTAATG |
| | Reverse Primer | 300 nM | CATCGTTCCTCTAAGAAGCTTTACAAT |
| | Probe | 200 nM | HEX/CCTTTTCCC/ZEN/TATTCTACTTAATTCGCAATTCATG |
| *ovale wallikeri* | Forward Primer | 300 nM | TTTTGAAGAATATATTAG-GATACATTATAG |
| | Reverse Primer | 300 nM | CATCGTTCCTCTAAGAAGCTTTACAAT |
| | Probe | 200 nM | FAM/CCTTTTCCCTTTTCTACTTAATTCGCTATTCATG/TAMRA |

Supplemental Table 2: PCR assay control reactions

| *Plasmodium* species | Parasites / µL | N Controls* | # Positive† | % Positive | Mean Ct | Standard deviation Ct |
|---|---|---|---|---|---|---|
| *malariae* | Std. 1 – 10,000 p/µL | 20 | 20 | 100 | 23.7 | 0.64 |
| | Std. 2 – 1000 p/µL | 20 | 20 | 100 | 27.4 | 0.69 |
| | Std. 3 – 100 p/µL | 20 | 20 | 100 | 31.1 | 0.96 |
| | Std. 4 – 10 p/µL | 20 | 20 | 100 | 34.5 | 1.44 |
| | Std. 5 – 1 p/µL | 20 | 6 | 30 | 37.4 | 1.47 |
| | Neg. – 0 p/µL | 20 | 0 | 0 | -- | -- |
| *ovale* spp. | Std. 1 – 10,000 p/µL | 19 | 19 | 100 | 27.7 | 2.93 |
| | Std. 2 – 1000 p/µL | 20 | 19 | 95 | 31.0 | 2.79 |
| | Std. 3 – 100 p/µL | 19 | 19 | 100 | 34.3 | 2.39 |
| | Std. 4 – 10 p/µL | 20 | 14 | *70* | 37.0 | 1.64 |
| | Std. 5 – 1 p/µL | 20 | 10 | 50 | 40.7 | 1.31 |
| | Neg. – 0 p/µL | 20 | 0 | 0 | -- | -- |
| *vivax* | Std. 1 – 10,000 p/µL | 20 | 20 | 100 | 25.7 | 0.54 |
| | Std. 2 – 1000 p/µL | 20 | 20 | 100 | 28.5 | 0.86 |
| | Std. 3 – 100 p/µL | 20 | 18 | 90 | 32.5 | 0.86 |
| | Std. 4 – 10 p/µL | 20 | 20 | 100 | 36.1 | 0.59 |
| | Std. 5 – 1 p/µL | 20 | 15 | 75 | 39.0 | 1.07 |
| | Neg. – 0 p/µL | 20 | 0 | 0 | -- | -- |
| *falciparum* | Std. 1 – 10,000 p/µL | 20 | 20 | 100 | 20.4 | 0.46 |
| | Std. 2 – 1000 p/µL | 20 | 20 | 100 | 24.4 | 0.29 |
| | Std. 3 – 100 p/µL | 20 | 20 | 100 | 27.7 | 0.35 |
| | Std. 4 – 10 p/µL | 20 | 19 | 95 | 31.7 | 0.26 |
| | Std. 5 – 1 p/µL | 20 | 10 | 50 | 35.5 | 0.84 |
| | Neg. – 0 p/µL | 20 | 0 | 0 | -- | -- |

Note: Negative controls used water for each *Plasmodium* species.
*Controls were run in duplicate per PCR plate.
†PCR assays in which negative controls amplified incorrectly (e.g., due to contamination) are re-run until controls remain negative; Results from original assays are then discarded as invalid and the re-run assay results are reflected in final analyses.

Supplemental Table 3: Comparison of characteristics between students in the full SMPS cohort vs. sample of students in the sub-cohort for non-falciparum analysis.

| Student characteristics | Full cohort | Sub-cohort | p-value* |
|---|---|---|---|
| n | 17,131 | 3,456 | |
| Age (years), median (IQR) | 11 (9, 13) | 11 (9, 13) | 0.884 |
| Male, n (%) | 8457 (49) | 1761 (51) | 0.093 |
| Malaria RDT+, n (%) | 3328 (20) | 686 (20) | 0.589 |
| Elevation (m), median (IQR) | 1230 (1058, 1458) | 1230 (1058, 1467) | 0.925 |
| Epidem. risk strata, n (%) | | | 0.966 |
| High | 8806 (51) | 1768 (51) | |
| Moderate | 2180 (13) | 448 (13) | |
| Low | 2952 (17) | 602 (17) | |
| Very low | 3193 (19) | 638 (18) | |
| Region, n (%) | | | 1 |
| Arusha | 2757 (16) | 552 (16) | |
| Iringa | 1583 (9) | 320 (9) | |
| Kagera | 3078 (18) | 619 (18) | |
| Mara | 2228 (13) | 452 (13) | |
| Mtwara | 1511 (9) | 307 (9) | |
| Rukwa | 1506 (9) | 301 (9) | |
| Tabora | 2045 (12) | 413 (12) | |
| Tanga | 2423 (14) | 492 (14) | |

IQR: Interquartile range: rapid diagnostic test
*Continuous variables are compared using a Kruskal-Wallis test. Categorical variables are compared using Chi-square test.

Supplement Table 4: Prevalences and 95% confidence intervals of species-specific malaria infections

| | Prevalence* | | |
|---|---|---|---|
| | n | % | 95% CI |
| Total student population | 3456 | 100.00 | --- |
| Total infections† | | | |
| Po | 814 | 23.6 | 22.2 – 25.0 |
| Pm | 136 | 3.9 | 3.3 – 4.6 |
| Pv | 11 | 0.3 | 0.2 – 0.6 |
| Pf | 755 | 21.8 | 20.5 – 23.3 |
| Single-species infections | | | |
| Po | 519 | 15.0 | 13.9 – 16.3 |
| Pm | 24 | 0.7 | 0.5 – 1.0 |
| Pv | 4 | 0.1 | 0.1 – 0.3 |
| Pf | 429 | 12.4 | 11.4 – 13.6 |
| Mixed-species infections‡ | | | |
| *P. falciparum co-infection* | | | |
| Po+Pf | 224 | 6.5 | 5.7 – 7.4 |
| Pm+Pf | 44 | 1.3 | 1.0 – 1.7 |
| Pv+Pf | 2 | 0.1 | 0.02 – 0.2 |
| Po+Pm+Pf | 55 | 1.6 | 1.2 – 2.1 |
| Po+Pv+Pf | 1 | 0.03 | 0.01 – 0.2 |
| Pm+Pv+Pf | 0 | 0 | 0 |
| Po+Pm+Pv+Pf | 0 | 0 | 0 |
| *Non-falciparum only* | | | |
| Po+Pm | 12 | 0.3 | 0.2 – 0.6 |
| Po+Pv | 3 | 0.1 | 0.03 – 0.3 |
| Pm+Pv | 1 | 0.03 | 0.01 – 0.2 |
| Po+Pm+Pv | 0 | 0 | 0 |

CI: Confidence interval; Pf: *Plasmodium falciparum;* Po: *Plasmodium ovale spp.*; Pm: *Plasmodium malariae*; Pv: *Plasmodium vivax*
*Prevalences represent point prevalences. Percents are calculated out of the total study population.
†Total infections include both single and mixed-species infections.
‡Categories are mutually exclusive.

Supplemental Table 5. Sensitivity analysis: *P. falciparum* and *P. malariae* prevalence estimated under a real-time PCR Ct of 45 (N=750)*,†

| *Plasmodium* species | Epidemiologic risk strata | | | Total | Crude prevalence | | Weighted prevalence§ | |
|---|---|---|---|---|---|---|---|---|
| | Low‡ n=250 | Moderate n=250 | High n=250 | N=750 | % | 95% CI | % | 95% CI |
| *P. falciparum* | | | | | | | | |
| PCR positive | 5 (2.0%) | 47 (18.8%) | 105 (42.0%) | 157 (20.9%) | 20.9% | 17.8%-24.5% | 24.6% | 20.6%-29.2% |
| Ct <40 | 5 (2.0%) | 45 (18.0%) | 103 (41.2%) | 153 (20.4%) | | | | |
| Ct 40 to <45 | 0 (0.0%) | 2 (0.8%) | 2 (0.8%) | 4 (0.5%) | | | | |
| PCR Negative | 245 (98.0%) | 203 (81.2%) | 145 (58.0%) | 593 (79.1%) | | | | |
| *P. malariae* | | | | | | | | |
| PCR positive | 2 (0.8%) | 6 (2.4%) | 12 (4.8%) | 20 (2.7%) | 2.7% | 1.6%-4.1% | 3.1% | 1.8%-4.9% |
| Ct <40 | 1 (0.4%) | 6 (2.4%) | 11 (4.4%) | 18 (2.4%) | | | | |
| Ct 40 to <45 | 1 (0.4%) | 0 (0.0%) | 1 (0.4%) | 2 (0.3%) | | | | |
| PCR Negative | 248 (99.2%) | 244 (97.6%) | 238 (95.2%) | 730 (97.3%) | | | | |

*Real-time PCR assay targeting the 18S ribosomal subunit; a Ct of 45 was employed for sensitivity analysis

†Sensitivity analysis population is a stratified random sample of the non-falciparum analysis population, where sampling was stratified by epidemiologic malaria risk strata to maintain malaria transmission heterogeneity within the sample.

‡Low epidemiological risk classification comprises ‘very low’ and ‘low’ strata.

§Weighted prevalence estimates are weighted to the distribution of students in the non-falciparum analysis population by epidemiologic malaria risk strata to account for oversampling of the moderate risk strata in the sensitivity analysis population.

Supplemental Table 6. Student infection characteristics by PCR+ vs. PCR– non-falciparum infection

| Student characteristics, n (%)*,† | *P. ovale* spp. | | | *P. malariae* | | | *P. vivax* | | |
|---|---|---|---|---|---|---|---|---|---|
| | PCR + | PCR - | | PCR+ | PCR - | | PCR+ | PCR - | |
| | Any Infection | No infection | p-value | Any Infection | No infection | p-value | Any Infection | No infection | p-value |
| N | 814 | 2138 | | 136 | 2138 | | 11 | 2138 | |
| Age (years) | | | | | | | | | |
| Median (IQR) | 11 (9, 13) | 11 (9, 13) | 0.128 | 12 (9, 13) | 11 (9, 13) | 0.038 | 11 (9.5, 12) | 11 (9, 13) | 0.95 |
| Male, n (%) | 402 (49.4) | 1075 (50.3) | 0.694 | 83 (61.0) | 1075 (50.3) | 0.019 | 8 (72.7) | 1075 (50.3) | 0.226 |
| Fever‡, n (%) | 23 (3.6) | 27 (1.5) | 0.003 | 12 (10.9) | 27 (1.5) | <0.001 | 0 (0.0) | 27 (1.5) | 1 |
| Epidemiological risk strata§, n (%) | | | <0.001 | | | <0.001 | | | 0.001 |
| High | 565 (69.4) | 770 (36.0) | | 125 (91.9) | 770 (36.0) | | 10 (90.9) | 770 (36.0) | |
| Moderate | 63 (7.7) | 327 (15.3) | | 10 (7.4) | 327 (15.3) | | 1 (9.1) | 327 (15.3) | |
| Low | 160 (19.7) | 433 (20.3) | | 1 (0.7) | 433 (20.3) | | 0 (0.0) | 433 (20.3) | |
| Very low | 26 (3.2) | 608 (28.4) | | 0 (0.0) | 608 (28.4) | | 0 (0.0) | 608 (28.4) | |

PCR: polymerase chain reaction; Po: *Plasmodium ovale spp.*; Pm: *Plasmodium malariae*; Pv: *Plasmodium vivax;* RDT: rapid diagnostic test

*Continuous variables are compared using a Kruskal-Wallis test. Categorical variables are compared using chi-square; fisher's exact test was applied where cell counts were <5.

†PCR negative represents students confirmed by PCR to have no malaria infection of any species.

‡Fever was defined as temperature at time of survey ≥38°C. Temperature is missing for n=618 (17.9%) subjects; percentages are calculated out of non-missing data.

§Epidemiological risk strata are defined as follows, based on *P.* falciparum prevalences in children from the 2014-15 Tanzanian SMPS: “very low” if prevalence <5%, “low” 5 to <10%, “moderate” 10 to <50%, and “high” ≥50%.
